# The Current State & Sentiment of Artificial Intelligence in North American Anesthesiology Residency Programs

**DOI:** 10.1101/2024.04.05.24305392

**Authors:** Tyler V Elliott, Joseph C Goldstein, Heidi V Goldstein

## Abstract

**Purpose:** This study aims to investigate the current state and sentiment of artificial intelligence (AI) training in North American anesthesiology residency programs, assessing existing AI education landscapes, identifying barriers to implementation, and understanding program directors’ expectations for AI’s impact on the field.

**Methods:** A cross-sectional survey targeted anesthesiology program directors across North America. The survey, conducted anonymously via Qualtrics, gauged their AI training offerings, sentiments towards AI’s influence, and familiarity with AI educational policies. Information on questionnaire development, administration, and data analysis was included.

**Results:** Of the 163 programs surveyed, 32 responded, yielding a response rate of 19.6%. A substantial 81% of responding program directors reported no current AI training. Despite this, 67% anticipate AI’s transformative impact. Only 19% currently offer AI/ML training. Standardized presentation of results with accompanying numerators and denominators were employed.

**Conclusion:** The findings reveal a significant gap between the recognition of AI’s importance and the current offering of training in anesthesiology residency programs. Barriers to implementation include resource constraints and time limitations, exacerbated by the pandemic. Overcoming these barriers and aligning positive sentiments with educational offerings is crucial for preparing future physicians for the AI-driven healthcare landscape.

**Implication Statement:** This study exposes a substantial gap between the positive anticipation of AI’s impact on anesthesiology and the current lack of training in North American residency programs. Recognizing this disparity is crucial for swiftly implementing comprehensive AI education, ensuring future anesthesiologists navigate healthcare’s evolving landscape adeptly.

**Disclosures:** The authors have no competing interests or financial interests to disclose. There was no funding associated with the completion of this study.

## Introduction

### Background & Significance

Artificial intelligence (AI) technology is rapidly becoming an integral part of our daily lives, poised to usher in profound changes across various domains, including healthcare. However, in today’s swiftly evolving healthcare landscape, AI is not merely a technological innovation; it is a significant transformational force that demands the attention of all health care professionals. The imperative of our time is not solely to acknowledge AI’s potential in medical care, but rather to recognize that every physician and trainee must possess a fundamental understanding of the technology. This literacy extends well beyond knowing the benefits of AI; it encompasses a deep grasp of its potential, its limitations, and the critical ethical and risk considerations that underpin its use. The crux of our mission is clear: we aim to empower medical professionals with the knowledge they need to drive positive change in the field of medicine and, most importantly, to advance patient care through a robust AI literacy.

By now it has been widely recognized that artificial intelligence (AI) technologies will increasingly impact the healthcare sector. In an official response, the Association of American Medical Colleges (AAMC) stated that AI “has tremendous potential to advance human health and usher in a new era of biomedicine [1].” The American Medical Association (AMA) also issued resolution 317-A-18, “Emerging Technologies (Robotics and AI) in Medical School Education,” calling for medical schools “to evaluate and update as appropriate their curriculum to increase students’ exposure to emerging technologies, in particular those related to robotics and artificial intelligence [2].” Multiple calls to action have been issued from leading organizations, including the National Academy of Medicine to “incorporate training in AI across health professions or risk creating a health workforce unprepared to leverage the promise of AI or navigate its potential perils [3].”

Historically, Anesthesiology has benefited greatly from advances in technology to improve patient safety and it is expected that AI will have a significant impact on this perioperative specialty [4]. Intelligent algorithms that can analyze large amounts of data can optimize preoperative risk modeling, intraoperative decision support, and pain management strategies [5-8]. Additionally, AI can assist in the prediction and early detection of adverse events, improving patient safety and outcomes when seconds count in the operating room.

### Objectives

Based on the anticipated major effect of AI on the specialty and the increasing realization that future anesthesiologists need to be AI literate [9], the authors were curious as to what the actual landscape of training and sentiment regarding artificial intelligence in North American anesthesiology residency programs looked like. With this study we hope to establish a baseline to guide leaders in the field in determining the needs and opportunities for an AI curriculum and related policies.

## Methods

### Research Question

“What is the current state and sentiment regarding artificial intelligence training in North American Anesthesiology residency programs?”

### Survey Design

This was a cross-sectional, anonymous survey conducted via the University of Florida’s Qualtrics survey software. It was aimed at anesthesiology residency program directors and/or designated faculty members responsible for training anesthesiology residents.

### Survey Instrument

The survey consisted of multiple-choice questions designed to gather information regarding the anesthesiology residency program’s state of artificial intelligence training, future plans, and the expected impact of AI on the specialty. The survey instrument underwent item generation, reduction, and formatting to gather information on AI training, future plans, and expected impacts. Pilot testing was conducted to ensure sensibility and reliability. The survey responses were anonymous and no identifying information was required when completing the survey.

### Sample Size and Selection Criteria

We contacted 168 anesthesiology residency program directors across North America, including the United States, Canada, and Mexico. The contact information for program directors was obtained online via publicly available information. All invited participants were contacted via email to respond to the survey.

### Data Collection and Analysis

The online survey was distributed via email to the anesthesiology residency program directors. The initial email contained a short description and link to the survey. Participants were given a four-week period to complete the survey. In an effort to combat response bias, two reminder emails were sent during this period. The survey was then closed, and the response data was analyzed using the Qualtrics survey software. Descriptive statistics were used to analyze the responses, with standardized presentation of results.

### Ethics

This survey was approved by the University of Florida’s Institutional Review Board (IRB #202201682) prior to distribution. All responses have remained completely anonymous as no identifying data was recorded. Before beginning the survey, informed consent was obtained from each participant. This included a detailed description of the survey’s focus, risks involved, explanation of anonymity, and contact information for the principal investigators as well as the university’s Institutional Review Board. All respondents were required to agree to participate before the survey would open, or they would be automatically directed to the end of the survey.

## Results

The survey was sent to 163 programs across the United States, Canada, and Mexico. There were 32 respondents to the survey, approximately a 19.6% overall response rate.

### Current Offering of AI/ML Training

A large majority, 26 out of 32 respondents (81%), reported no current AI/ML training.

### Sentiment Regarding AI’s Impact

A similar majority, 27 out of 32 respondents (84%), anticipate AI’s transformative impact.

### Implementation of AI/ML Training

Only 6 out of 32 respondents (19%) currently offer AI/ML fundamental training.

### Barriers to Implementation

Various barriers were cited, including lack of time, funding, and faculty expertise.

### Familiarity with AI Medical Educational Policy

Only 1 out of 27 respondents reported being familiar with recent AI educational policy changes.

## Discussion

To our knowledge, this study represents the first attempt to assess the current state and sentiment regarding AI incorporation within North American anesthesiology residency programs. While the majority of respondents express the importance of integrating AI training into their programs and foresee AI bringing transformative changes to the specialty, less than one-fifth currently provide any related training. These findings underscore a significant gap between the recognized importance of AI and the current ability to offer relevant training. Moreover, various barriers identified by respondents, including the lack of qualified faculty, support, and time within the curriculum, further accentuate this gap. It is our hope that these insights will assist specialty leaders in preparing their programs for the ongoing integration of artificial intelligence in medicine.

The study’s findings reveal a notable discord between the optimistic expectations of program directors regarding AI’s transformative impact on anesthesiology and the current state of AI education in residency programs across North America. While a significant proportion anticipate AI’s positive influence, an overwhelming majority of programs do not offer any AI/ML training at present. Despite this scarcity, a substantial portion of program directors foresee significant changes in anesthesia due to AI, with many even categorizing AI as a potential disruptor to the field.

This optimism, however, contrasts sharply with the insufficient availability of introductory AI/ML training, highlighting the urgent need for alignment between positive sentiments and educational offerings. A sustained and comprehensive approach to training implementation is crucial, as evidenced by the preference among respondents for incorporating AI/ML curricula through a series of educational sessions rather than one-off experiences. Such an approach enhances residents’ understanding and practical application of AI concepts, better preparing them for the evolving landscape of AI in healthcare.

Barriers to implementation, echoing familiar challenges in medical education, such as resource constraints and time limitations, may disproportionately affect smaller programs, underscoring the need for targeted efforts to overcome these obstacles and promote widespread integration of an AI-focused education.

However, several limitations temper the interpretation of our survey findings. The anonymous survey format obscured specific geographic distributions among respondents, hindering the assessment of regional variations in AI incorporation. Additionally, lacking details on program sizes limits our insight into potential differences in their capacity to integrate AI training. While larger, more established programs may possess greater resources, smaller programs could face constraints. The use of unsolicited email contacts as a distribution method introduces potential biases and may have influenced the response rate. While typical for email surveys, caution is warranted due to potential variations and non-response bias. A more official outreach could have improved participation and captured a more representative sample of anesthesiology residency programs. Moreover, our cross-sectional study provides a snapshot rather than a dynamic, longitudinal view of the evolving integration of AI. It lacks the ability to track changes over time.

To address these limitations, targeted outreach efforts can be undertaken to engage program directors from underrepresented regions or smaller programs, ensuring a more diverse and representative sample. Collaborative efforts with professional organizations could facilitate access to a broader pool of participants and enhance the generalizability of findings. Furthermore, longitudinal studies tracking changes in AI training offerings and sentiments over time could provide valuable insights into the evolving landscape of AI education in anesthesiology.

Despite these limitations, our data underscores the perceived importance of integrating AI fundamentals into the anesthesiology residency curriculum and serves as a starting point for future studies. This resonates with the broader recognition of AI’s transformative potential in healthcare, emphasizing the need to prepare future physicians for this evolving landscape.

## Conclusion

In summary, this study highlights a significant gap between the recognized importance of AI in anesthesiology and the current availability of AI training within North American residency programs. Addressing barriers such as faculty expertise and resource constraints is essential to prepare future anesthesiologists for the evolving healthcare landscape. By fostering comprehensive AI education, we can ensure that residents are equipped to leverage AI effectively, ultimately advancing patient care in the era of artificial intelligence.

**Figure 1.**
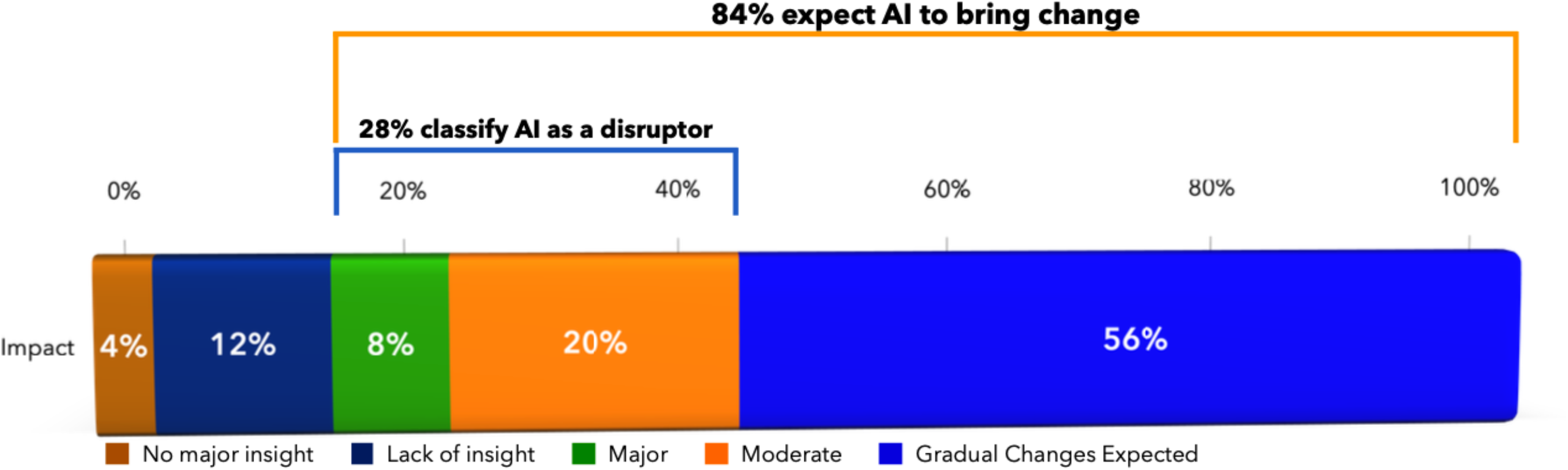

**Figure 2.**
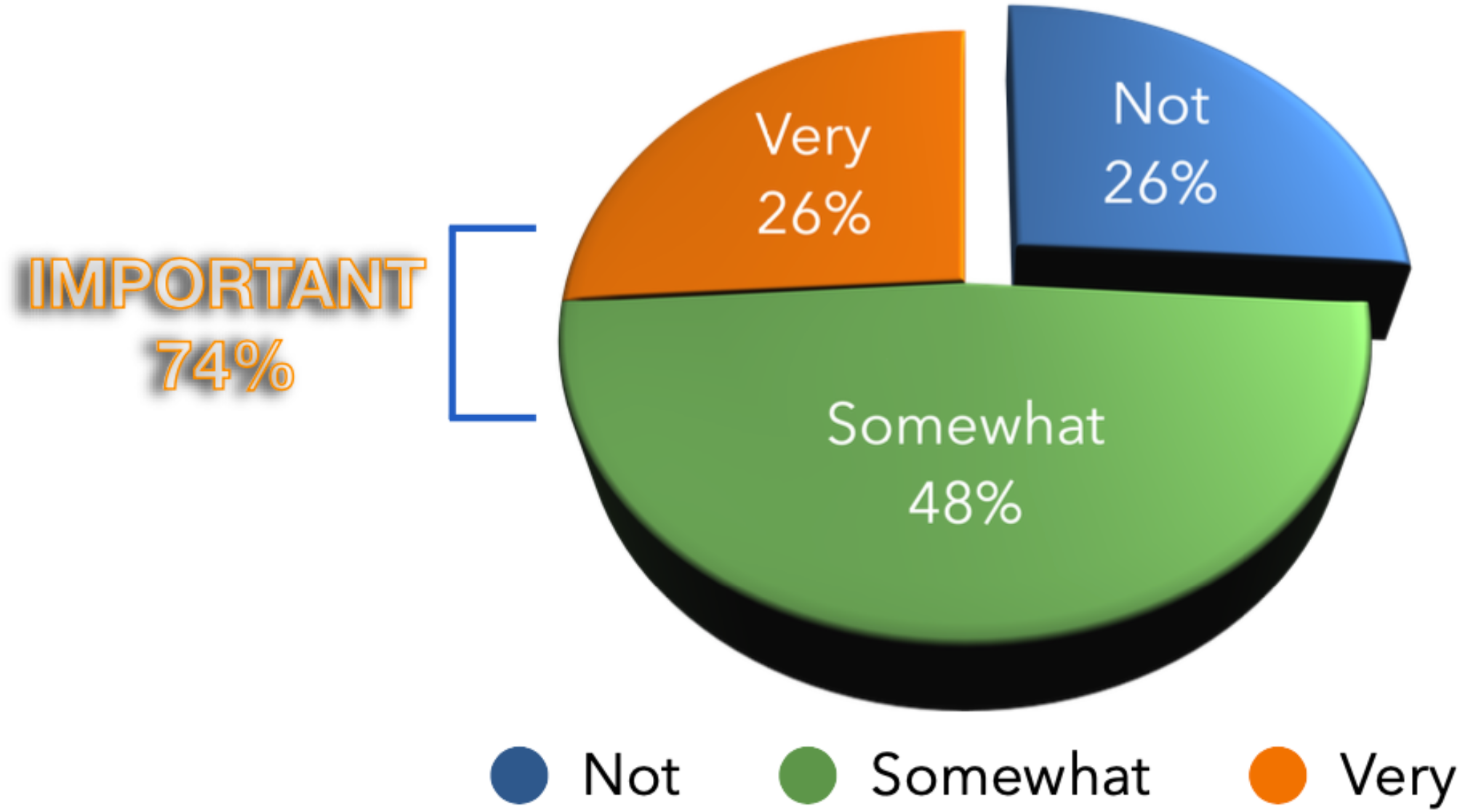

## Notes

### Competing Interest Statement

The authors have declared no competing interest.

### Funding Statement

This study did not receive any funding

### Author Declarations

Ethics committee/IRB of University of Florida waived ethical approval for this work

